# The Association between Mental Health and Cognitive Ability: Evidence from The UK Household Longitudinal Study

**DOI:** 10.1101/2025.01.26.25321154

**Authors:** Syka Iqbal, Muhammad Waqas, Min Hooi Yong, Ronan McGarrigle, Eleftheria Vaportzis

## Abstract

The relationship between poor mental health and cognitive impairments in older age is well-established. Social engagement also influences cognitive ability; however much of the research on this relationship has not accounted for the impact of mental health and demographic factors. This study examined the associations between cognitive ability and mental health in older adults, controlling for social interaction and socio-demographic factors. In total, 7,685 individuals aged 65 or older were drawn from the UK Household Longitudinal Study Understanding Society (1). Cognitive abilities were assessed using self-reports and performance on five tasks (immediate and delayed word recall, subtraction, number series and numerical ability). Mental health scores were derived from the General Health Questionnaire (GHQ-12). We controlled for social interaction, gender, ethnicity, educational background, marital status, number of children, and geographic location. We found positive relationships between mental health and all measures of cognitive ability except performance on subtraction and number series tasks. These relationships remained after controlling for social interaction. Demographic factors that contributed to the relationship between mental health and cognitive ability included being White, having higher education, being male for numerical tasks only, and being female, married or divorced for verbal memory tasks only. Overall, our results suggest that the relationship between mental health and cognitive abilities persists when controlling for social interaction alongside socio- demographic factors in older adults, underscoring the importance of addressing these factors in policies and interventions for healthy ageing.

## 1. Introduction

Older adulthood is a time of multiple changes in the cognitive, functional and social domains (2, 3). In particular, older adults are shown to be at increased risk of cognitive decline, loneliness, poor health and socio-economic deprivation (4), and these factors are associated with poor mental health outcomes in older adults such as depression and dementia (5). There is a need for better understanding the determinants of cognitive decline and poor mental health.

Providing appropriate care for older adults is a global challenge. With longevity increasing steadily, there is a pressing need to better understand ageing populations and the relationship between cognitive decline and mental health. Maintaining cognitive abilities into old age is crucial for coping with chronic disease symptoms, quality of life, performing self-care, adhering to medication instructions and promoting independence in older adults (6).

Although cognitive function is a concern for older adults, cognitive decline is not inevitable in healthy ageing with research suggesting different domains decline at different rates (7). For example, cognitive abilities such as vocabulary are resilient to the ageing process and may even improve gradually over time (8).

Baumeister and Bunce (2015) found that the relationship between mental health and cognitive decline varied within individuals, affecting certain cognitive functions more than others. For example, there were significant impairment in executive function and processing speed, while domains like vocabulary remained stable. This within-person variability suggests that cognitive decline in older adults is selective with specific functions potentially impacted more than others by mental health changes.

Numerous studies have established a correlation between poor mental health and cognitive impairments in older adulthood. Gallagher, Kiss (9) conducted a study involving 7,610 older adults, with 1,133 (14.9%) of which reported experiencing clinically significant depressive symptoms at the baseline and exhibited pronounced cognitive impairments. The study specifically examined older adults, so no direct comparisons were made with younger populations. Despite evidence of the relationship between mental health and cognitive decline in recent years, a gap remains in understanding specific cognitive functions that may be impacted by declining mental health. These gaps are further compounded by the ambiguity surrounding the relationship between depression and cognitive outcomes. For example, depression has been shown to negatively impact specific cognitive domains, such as episodic memory, executive function, working memory and processing speed (McDermott & Ebmeier, 2009, Haile 2022, Maksyutynska et al, 2024). However, not all research has been consistent with regard to the effects on isolated cognitive functions. For example, research showed that rates of depression were negatively correlated with incidental memory, executive function (cognitive processes such as planning and problem solving) and overall functionality (the ability to perform tasks and daily activities) (Costa Dias, 2017, Wang et al, 2017), highlighting the nuanced relationship between mental health and cognition.

The relationship between mental health and cognitive decline may also be related to social network size and feelings of loneliness. A lack of significant connections and social integration is associated with barriers to visiting friends and fosters increased feelings of loneliness (Victor et al., 2000). In a longitudinal study of 3,777 older individuals assessed at baseline, it was found that those who received more support throughout their lifetime, irrespective of the type of support they received, had a 55% reduced risk of dementia and a 53% reduced risk of Alzheimer’s disease (8). This indicates that strong social networks and a robust support system are critical for mental health in older adults. Furthermore, a qualitative study involving 59 participants found the central component of loneliness is a lack of connection with others (10). Harada (8) found the relationship between loneliness and mental health was bi-directional; mental health conditions reduced the capacity to socialise with others and led to withdrawal from social activities. On the other hand, reduced socialisation led to a decline in mental health which precipitated further isolation, compounding the downward spiral. Social networks and loneliness are a complex situation in which there are many possible factors that could influence the degree of loneliness in individuals, but having friends are a known variable that minimises feelings of loneliness thus improving mental health.

Yet, much of the research on loneliness and reduced cognitive ability has not accounted for mental health impacts and demographic factors. Seshadri, Wolf (11) estimated the ‘remaining lifetime risk’ of developing Alzheimer’s disease in individuals aged 65 revealing a substantial gender disparity, with females exhibiting a twofold higher risk (12%) in comparison to males (6.3%). Additionally, Kuiper, Smidt (12) examined the associations between social network size, loneliness, and cognitive performance using neuropsychological tests measuring four cognitive domain scores of processing speed, interference control, verbal memory, and working memory in a sample of 378 depressed older adults. The study identified a moderate negative relationship between loneliness and working memory capacity. In another study, lower-educated and lower income depressed older adults were found to have an increased risk of loneliness (13). Reduced opportunities for social interaction may have contributed to this association. Other research has highlighted context-specific factors (e.g., number of children) to reflect changing demographics, including declining fertility rates. For example, Ajrouch, et al. (2017) highlighted the central role of children in providing informal support to older adults. Other important demographic factors to consider include education, gender and number of children, as part of a complex system that more accurately captures the complex relationship between cognition and mental health (14), Wu et al, 2023).

The current study aimed to examine the associations between cognitive ability and mental health. We employed a series of cognitive tests in a large nationally representative sample of older people in the UK, using Wave 3 United Kingdom Health Longitudinal Survey data (UKHLS) to provide insights into the specific cognitive functions associated with poor mental health in older age. This dataset allows us to conduct a highly-powered statistical analysis and control for social interaction and socio-demographic factors such as gender, ethnicity, educational background, marital status, and number of children, to investigate the relationships between mental health, cognitive function, and social interaction in older adulthood. Rather than addressing single factors, this exploratory study aims to shed light on the intricate relationship between mental health and cognitive decline in older adulthood.

## 2. Materials and Methods

We completed secondary analysis of de-identified cross-sectional data from Wave 3 of the UK Household Longitudinal Study (UKHLS; 15). We obtained the data from the Understanding Society website (www.understandingsociety.ac.uk). In this paper, we summarise key aspects of the development of the survey and its methodology; a complete account can be obtained from several other reports (see Buck & McFall, 2011; McFall, 2012; McFall & Garrington, 2011).

### 2.1. Participants

UKHLS is a longitudinal study interviewing everyone in a household to see how different generations experience life in the UK. The first wave of data collection was completed between January 2009 and December 2011. Sampling from the Postcode Address File in Great Britain and the Land and Property Services Agency list of domestic properties in Northern Ireland identified 55,684 eligible households. Interviews were completed with a total of 50,994 individuals aged 16 or older from 30,117 households. At Wave 3, interviews were completed with a total of 49,768 individuals aged 16 or older from 27,715 households (McFall, 2012a). The current study uses the data of 7,685 individuals aged 65 or older from Wave 3 only; their demographic data are summarised in Table 1.

**Table 1.** Demographic characteristics of 7,685 participants over 65 years in Wave 3.

| **Demographic** | **N** | **%** | **SD** |
| --- | --- | --- | --- |
| Gender |  |  |  |
| Female | 4,158 | 54.1 | 0.50 |
| Male | 3,527 | 45.9 | 0.50 |
| Ethnicity |  |  |  |
| White | 7,362 | 95.8 | 0.20 |
| Other | 323 | 4.2 | 0.20 |
| Education |  |  |  |
| University degree | 1,821 | 23.7 | 0.43 |
| Other qualification | 3,328 | 43.3 | 0.50 |
| No qualification | 2,498 | 32.5 | 0.47 |
| Marital status |  |  |  |
| Married | 4,795 | 62.4 | 0.48 |
| Divorced | 699 | 9.1 | 0.29 |
| Widowed | 1,798 | 23.4 | 0.42 |
| Single | 392 | 5.1 | 0.05 |
| Children living in the household |  |  |  |
| None | 7,654 | 99.6 | 0.06 |
| One | 23 | 0.3 | 0.05 |
| Two | 8 | 0.1 | 0.02 |
| Location |  |  |  |
| England | 5,687 | 74 | 0.44 |
| Wales | 730 | 9.5 | 0.29 |
| Scotland | 784 | 10.2 | 0.30 |
| Northern Ireland | 484 | 6.3 | 0.24 |

166 Note. Values may not add up due 100% due to rounding or missing values.

### 2.2. Procedures

Data collection was primarily undertaken using Computer Assisted Personal Interviewing, including self-completion (CAPI).

### 2.3. Measures

#### 2.3.1. Cognitive assessment

Participants were asked to self-report on their own cognitive ability. The following question was asked, “First, how would you rate your memory at the present time? Would you say it is excellent, very good, good, fair or poor?” Moreover, five cognitive function tests were used in this study and are reported. Self-reported cognitive ability measure score ranges from 1 to 5 and all other cognitive ability measures’ scores range from 0 to 100.

#### 2.3.2. Immediate and delayed word recall

Participants listened to a list of ten words delivered by a computer. They were asked to immediately recall the words, and then again at a later stage without having heard them again. Correct responses of a maximum score of ten were recorded on each test.

#### 2.3.3. Subtraction

Participants were asked to subtract 7 from 100 and then subtract 7 again from their answer four more times. Correct responses of a maximum of five attempts were recorded.

#### 2.3.4. Number series

Participants were presented with a number sequence in which they populated the gaps in a logical series. Participants were administered two sets of three number sequences. The difficulty of the second set was determined by participants’ performance on the first set. A total score was derived accounting for the difficulty of the items.

#### 2.3.5. Numerical ability

Participants were given six numerical problems to solve. For example, the first question asked “In a sale, a shop is selling all items at half price. Before the sale, a sofa costs £300. How much will it cost in the sale?”. Based on participants’ responses they were administered either one simpler additional problem or two more difficult problems. Correct responses were recorded.

#### 2.3.6. Mental health

General Health Questionnaire (GHQ-12; 16). The GHQ-12 is a validated screening measure of risk of mental health issues (17). It comprises 12 items concerning symptoms about general happiness, confidence, ability to face problems, make decisions, overcome difficulties, and enjoy day-to-day activities over the past four weeks. Six items are worded positively and six are worded negatively. Participants self-reported their symptoms using a four-point Likert scale relating to the frequency or severity of the symptom in comparison to what is usual for the respondent (e.g., 1 = more than usual, 2 = about the same as usual, 3 = less than usual, 4 = much less than usual). Scores can range between 0 to 36 with 36 representing the lowest level of subjective well-being. The higher the score, the more likely it is that respondents are suffering from some form of psychological distress. For ease of interpretation, we reversed the overall score so that a value of 36 represents the highest level and going forward we refer to this variable simply as mental health.

#### 2.3.7. Social Interaction

Social interaction was measured by the question ’Do you go out socially or visit friends when you feel like it?’. The answer to this question is binary (Yes, No).

### 2.4. Ethical approval

UKHLS is designed and conducted in accordance with the ESRC Research Ethics Framework and the ISER Code of Ethics. The University of Essex Ethics Committee approved Wave 3 of UKHLS. The project received ethical approval at Wave 1 by the National Research Ethics Service (NRES) Oxfordshire REC A (08/H0604/124), at BHPS

Wave 18 by the NRES Royal Free Hospital & Medical School (08/H0720/60) and at Wave 4 by NRES Southampton REC A (11/SC/0274).

## 3. Results

### 3.1. Descriptive statistics

A total of 7,685 participants from Wave 3 that met the inclusion criteria were included in the analyses.

### 3.2. Cognition and mental health

Table 2 shows the Ordinary Least Square (OLS) regression coefficients for all cognitive assessment scores. For the regression analyses, we used a cognition variable as an outcome with socio-demographic variables (gender, ethnicity, educational levels, marital status, and number of children) as covariate and GHQ scores as a predictor. We applied this analysis to self-report cognitive ability and for each of the five cognitive tasks.

**Table 2.** Regression analyses with cognitive tasks as outcome, GHQ as predictor and socio- demographic variables as covariates.

| Variables | (1) | (2) | (3) | (4) | (5) | (6) |
| --- | --- | --- | --- | --- | --- | --- |
|  | Self-report<br>Cognitive<br>Ability | Immediate<br>Word<br>Recall | Delayed<br>Word<br>Recall | Numerical<br>Ability | Subtraction | Number<br>Series |
| GHQ | 0.05***<br>(0.00) | 0.34***<br>(0.00) | 0.33***<br>(0.00) | 0.31***<br>(0.00) | 0.26**<br>(0.00) | 0.08<br>(0.18) |
| Male | -0.14***<br>(0.00) | -5.53***<br>(0.00) | -5.96***<br>(0.00) | 4.54***<br>(0.00) | 5.43***<br>(0.00) | 3.52***<br>(0.00) |
| White | 0.13*<br>(0.02) | 7.02***<br>(0.00) | 7.61***<br>(0.00) | 13.39***<br>(0.00) | 6.10**<br>(0.00) | 3.05<br>(0.05) |
| Higher edu | 0.21*** | 11.85*** | 12.12*** | 15.38*** | 13.68*** | 10.44*** |
|  | (0.00) | (0.00) | (0.00) | (0.00) | (0.00) | (0.00) |
| Other edu | 0.10*** | 6.67*** | 7.32*** | 9.55*** | 8.27*** | 2.39*** |
|  | (0.00) | (0.00) | (0.00) | (0.00) | (0.00) | (0.00) |
| Married | -0.05 | 3.76*** | 4.20*** | 4.06*** | -0.75 | 1.78 |
|  | (0.36) | (0.00) | (0.00) | (0.00) | (0.66) | (0.20) |
| Divorced | 0.00 | 2.98** | 3.10* | 1.60 | -1.25 | 0.05 |
|  | (0.94) | (0.00) | (0.01) | (0.19) | (0.54) | (0.98) |
| Widowed | -0.02 | -1.59 | -1.97 | -0.22 | -1.76 | -0.35 |
|  | (0.75) | (0.09) | (0.07) | (0.84) | (0.34) | (0.81) |
| 1 child | -0.10 | 3.72 | 5.52* | 3.47 | -2.33 | 3.56 |
|  | (0.62) | (0.12) | (0.04) | (0.27) | (0.75) | (0.64) |
| >1 child | 0.31 | 5.85 | 13.55** | -12.85*** | 13.27*** | 11.99 |
|  | (0.57) | (0.20) | (0.01) | (0.00) | (0.00) | (0.58) |
| 2.country | -0.01 | -1.04 | -0.98 | -1.46* | -0.29 | 0.25 |
|  | (0.87) | (0.11) | (0.19) | (0.04) | (0.82) | (0.82) |
| 3.country | -0.06 | -1.89** | -1.25 | -1.67* | 4.18*** | 0.69 |
|  | (0.10) | (0.00) | (0.09) | (0.02) | (0.00) | (0.51) |
| 4.country | -0.11* | -1.64* | 0.44 | 1.70 | 3.80* | 3.84* |
|  | (0.02) | (0.04) | (0.64) | (0.08) | (0.02) | (0.01) |
| Constant | 1.58*** | 32.16*** | 18.22*** | 42.60*** | 58.11*** | -4.40 |
|  | (0.00) | (0.00) | (0.00) | (0.00) | (0.00) | (0.09) |
| Observations | 7,685 | 7,619 | 7,616 | 7,600 | 7,385 | 6,967 |
| F | 38.80 | 78.36 | 67.70 | 102.38 | 34.57 | 14.04 |
| R<sup>2</sup> | 0.06 | 0.12 | 0.10 | 0.17 | 0.04 | 0.03 |

Note: Gender (1 – Male, 2 – Female), Edu – education. GHQ – general health questionnaire.
\*\*\* $p < .001$ , \*\* $p < .01$ , \* $p < .05$ .

### 3.3. Self-reported cognitive ability

Results showed that the model was significant with good mental health and the following covariates (female, White, higher educational levels) as significant predictors, F (13, 7671) = 38.80, R^2^ = .063, *p* < .001. See Model 1. Specifically, results showed that having good mental health is positively associated with higher self-reported cognitive ability (coef = 0.05, *p* < .001). In terms of the covariate results, being female compared to males, White compared to non-Whites and those with higher education were also significant predictors associated with self-reported cognitive ability, all *p*s < .02. See Table 2 for full details. Marital status and number of children at home were not significant predictors (all *p*s > .36).

### 3.4. Individual cognitive tasks

We have five individual cognitive tasks: immediate word recall, delayed word recall, numerical ability, subtraction and number series that we have identified as an outcome.

Our results showed similar trends for immediate word recall (Model 2), delayed word recall (Model 3), numerical ability (Model 4), subtraction (Model 5), in that a higher GHQ score was positively associated with better cognitive outcome, all *p*s < .001. Unlike the other cognitive outcomes, having higher GHQ did not significantly predict number series (Model 6) outcome (coef = 0.08, *p* = 0.18), R^2^ = 0.03.

In addition to being female, White, and highly educated , being married or divorced compared to singles was also a significant predictor for immediate and delayed word recall, and numerical ability (married and divorced performed better; all *p*s < .001; see Model 2 and 3). For the numerical ability and subtraction outcomes (Model 4 and 5), being male compared to females were a significant predictor, both *p*s < .001. Other demographics such as being White and having higher education were also significant predictors for Model 2 to 5 (those being White and having higher education performed better; all *p*s < .001).

### 3.5. Self-reported cognition, mental health, social interaction

We added the *social interaction* variable as a predictor to the regression analyses to determine whether changes in cognitive ability were associated with mental health, social interaction with friends. Socio-demographic variables including gender, ethnicity, educational background, marital status, and number of children, were added as covariates. The regression analyses revealed that having a higher GHQ (coef = 0.04, *p* < .001) and more social interaction with their friends (coef = 0.07, *p* =.02) were significant predictors of overall cognitive ability, F (14, 7666) = 36.69, R^2^ = 0.06, *p* < .001 (see Model 7 in Table 3). In terms of covariates, being female compared to males, White compared to non-White, and having higher education levels compared to lower education were all significant predictors associated with this model, all *p*s < .02. Marital status and number of children in the household were not significant predictors to this model, all *p*s > .31.

**Table 3.**
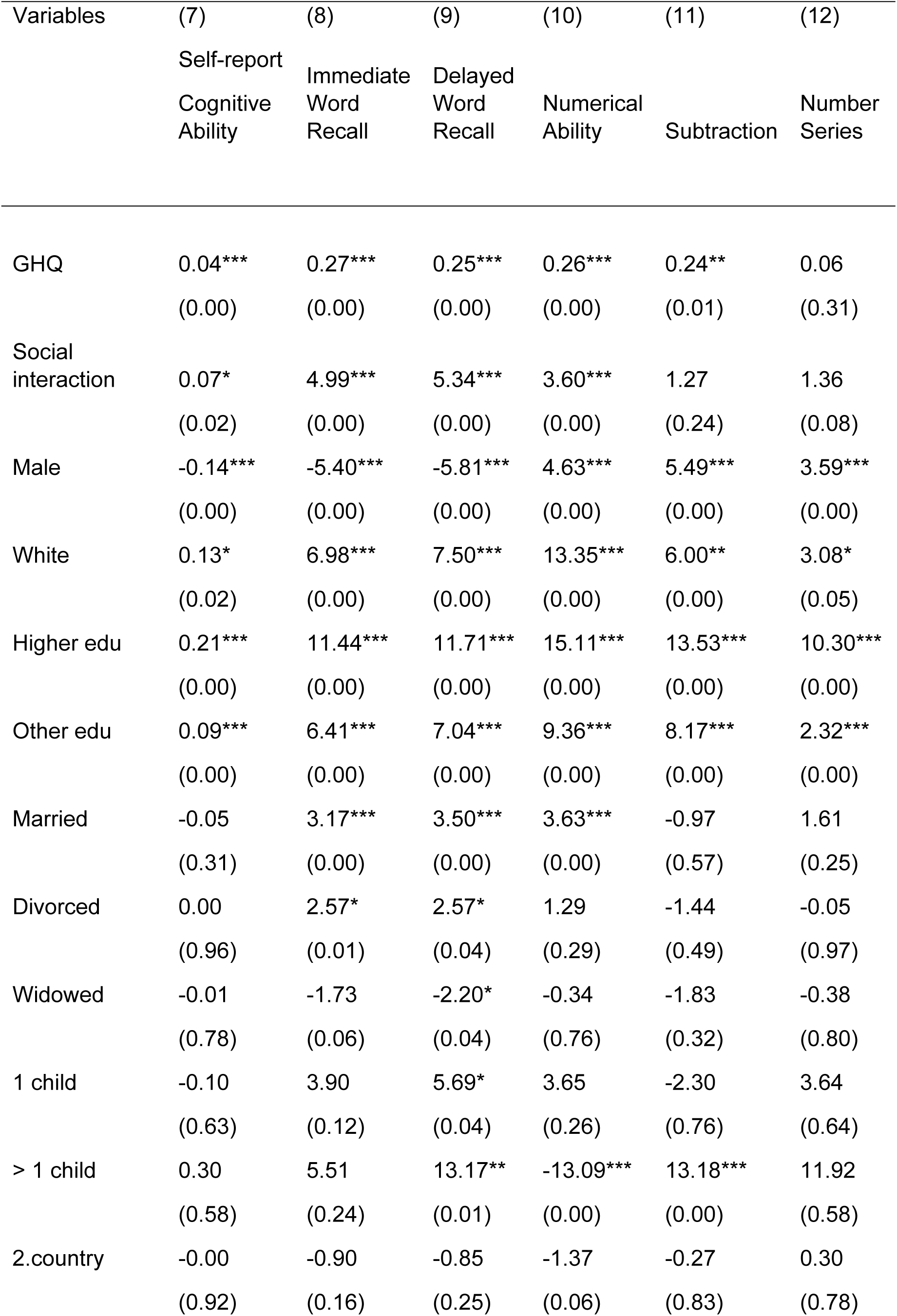
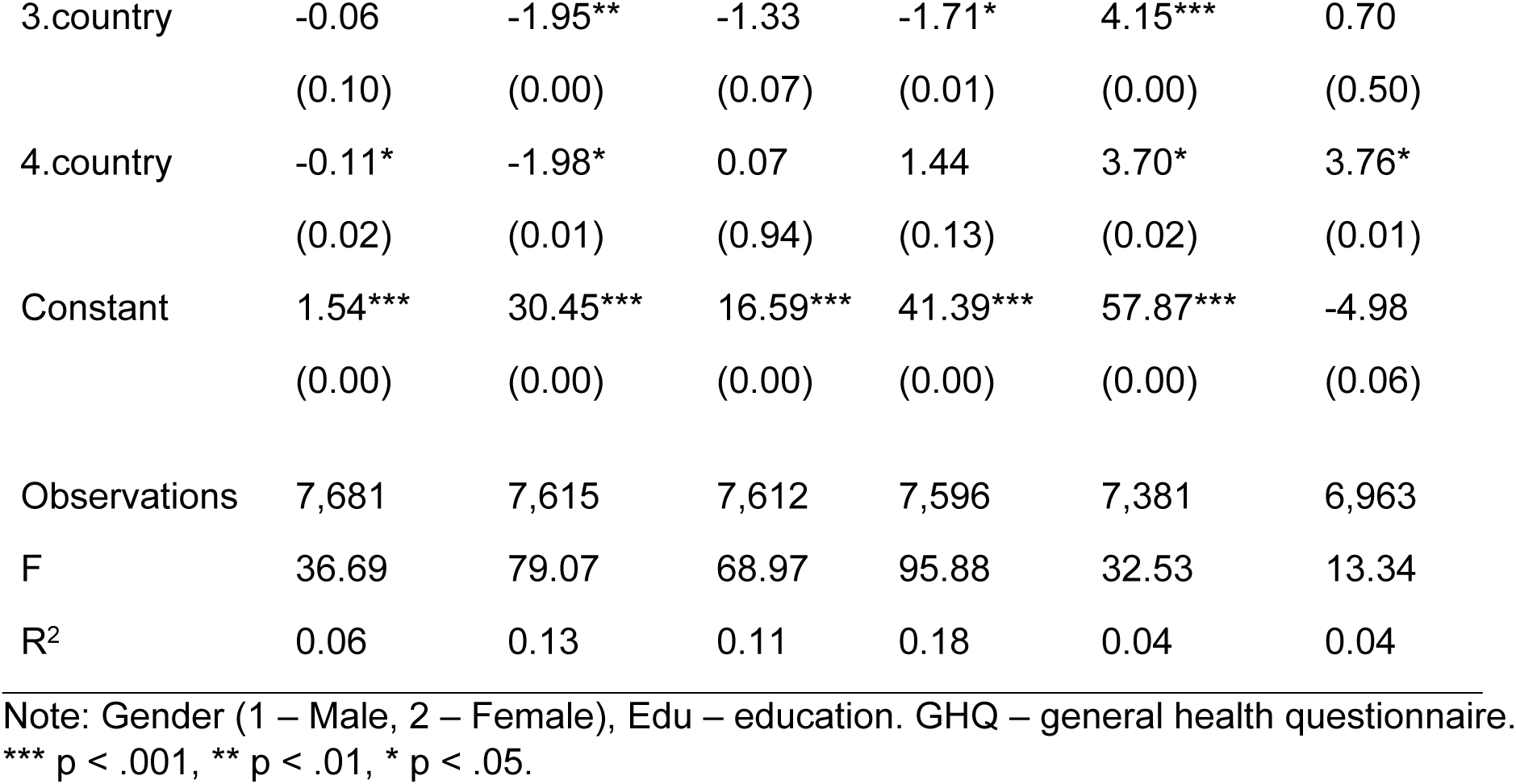
Regression analyses with cognitive tasks as outcome, GHQ as predictor, and social interaction and socio-demographic variables as covariates.

| Variables | (7) | (8) | (9) | (10) | (11) | (12) |
| --- | --- | --- | --- | --- | --- | --- |
|  | Self-report<br>Cognitive<br>Ability | Immediate<br>Word<br>Recall | Delayed<br>Word<br>Recall | Numerical<br>Ability | Subtraction | Number<br>Series |
| GHQ | 0.04***<br>(0.00) | 0.27***<br>(0.00) | 0.25***<br>(0.00) | 0.26***<br>(0.00) | 0.24**<br>(0.01) | 0.06<br>(0.31) |
| Social<br>interaction | 0.07*<br>(0.02) | 4.99***<br>(0.00) | 5.34***<br>(0.00) | 3.60***<br>(0.00) | 1.27<br>(0.24) | 1.36<br>(0.08) |
| Male | -0.14***<br>(0.00) | -5.40***<br>(0.00) | -5.81***<br>(0.00) | 4.63***<br>(0.00) | 5.49***<br>(0.00) | 3.59***<br>(0.00) |
| White | 0.13*<br>(0.02) | 6.98***<br>(0.00) | 7.50***<br>(0.00) | 13.35***<br>(0.00) | 6.00**<br>(0.00) | 3.08*<br>(0.05) |
| Higher edu | 0.21***<br>(0.00) | 11.44***<br>(0.00) | 11.71***<br>(0.00) | 15.11***<br>(0.00) | 13.53***<br>(0.00) | 10.30***<br>(0.00) |
| Other edu | 0.09***<br>(0.00) | 6.41***<br>(0.00) | 7.04***<br>(0.00) | 9.36***<br>(0.00) | 8.17***<br>(0.00) | 2.32***<br>(0.00) |
| Married | -0.05<br>(0.31) | 3.17***<br>(0.00) | 3.50***<br>(0.00) | 3.63***<br>(0.00) | -0.97<br>(0.57) | 1.61<br>(0.25) |
| Divorced | 0.00<br>(0.96) | 2.57*<br>(0.01) | 2.57*<br>(0.04) | 1.29<br>(0.29) | -1.44<br>(0.49) | -0.05<br>(0.97) |
| Widowed | -0.01<br>(0.78) | -1.73<br>(0.06) | -2.20*<br>(0.04) | -0.34<br>(0.76) | -1.83<br>(0.32) | -0.38<br>(0.80) |
| 1 child | -0.10<br>(0.63) | 3.90<br>(0.12) | 5.69*<br>(0.04) | 3.65<br>(0.26) | -2.30<br>(0.76) | 3.64<br>(0.64) |
| > 1 child | 0.30<br>(0.58) | 5.51<br>(0.24) | 13.17**<br>(0.01) | -13.09***<br>(0.00) | 13.18***<br>(0.00) | 11.92<br>(0.58) |
| 2.country | -0.00<br>(0.92) | -0.90<br>(0.16) | -0.85<br>(0.25) | -1.37<br>(0.06) | -0.27<br>(0.83) | 0.30<br>(0.78) |
| 3.country | -0.06<br>(0.10) | -1.95**<br>(0.00) | -1.33<br>(0.07) | -1.71*<br>(0.01) | 4.15***<br>(0.00) | 0.70<br>(0.50) |
| 4.country | -0.11*<br>(0.02) | -1.98*<br>(0.01) | 0.07<br>(0.94) | 1.44<br>(0.13) | 3.70*<br>(0.02) | 3.76*<br>(0.01) |
| Constant | 1.54***<br>(0.00) | 30.45***<br>(0.00) | 16.59***<br>(0.00) | 41.39***<br>(0.00) | 57.87***<br>(0.00) | -4.98<br>(0.06) |
| Observations | 7,681 | 7,615 | 7,612 | 7,596 | 7,381 | 6,963 |
| F | 36.69 | 79.07 | 68.97 | 95.88 | 32.53 | 13.34 |
| R<sup>2</sup> | 0.06 | 0.13 | 0.11 | 0.18 | 0.04 | 0.04 |

Note: Gender (1 – Male, 2 – Female), Edu – education. GHQ – general health questionnaire.
\*\*\* p < .001, \*\* p < .01, \* p < .05.

### 3.6. Individual cognitive tasks, mental health, social interaction

Similar to the earlier Results section, we have five individual cognitive tasks as an outcome. When we reviewed the regression analyses with individual cognitive tasks, results were similar to the overall self-report cognitive ability for immediate (Model 8) and delayed word recall (Model 9) and numerical ability (Model 10) in that both having higher GHQ and more interaction with friends were significant predictors of all cognitive abilities, all *p*s < .001 .

Similar significant covariates (female, White, higher education) were also significant predictors in Models 8 and 9, all ps < .001. Compared to singles, being married, both *p*s < .001, or divorced, both *p*s < .004 were also significant predictors in Models 8 and 9. However for Model 10, being male compared to female was a significant predictor in this model, *p* < .001 as well as being White, highly educated and married, all *p*s < .001. For Model 11 with subtraction as a cognitive outcome, the regression analyses showed that having a higher GHQ was a significant predictor (coef = 0.24, *p* =.01), but not for social interaction (coef = 1.27, *p* =.24) in this model, R^2^ = 0.04. Being male, White and having higher education were also significant predictors in the model 11, all *p*s < .001. Results for Model 12 (number series as an outcome) showed that GHQ and social interaction were both not significant predictors, both *p*s > .08.

## 4. Discussion

We explored associations between cognitive ability and mental health in a large representative population-based sample of over 7,000 individuals aged 65 or older. We observed associations between mental health, self-reported cognitive ability, and cognitive ability as measured by the immediate word recall, delayed word recall, numerical ability, and subtractions tasks. Better mental health was associated with better cognitive outcomes as a whole. These results confirm those reported previously (18).

We found positive associations between mental health and all cognitive tasks for those who identified themselves as White compared to non-White and those with higher education compared to those with lower education levels. Studies that investigated mental health and cognition separately have shown differences due to ethnicity and educational attainment.

Native-born Whites had significantly better mental health outcomes compared to native-born Blacks (19). Non-Hispanic Whites had significantly better cognitive scores compared to Hispanics and Non-Hispanic Blacks (20). Results from a large sample of 502,357 individuals from the UK Biobank cohort demonstrated that higher education was associated with better cognitive performance (21). Higher educational achievement was also associated with better mental health outcomes in older age (22). Taken together, it appears that having higher education serves as a protective factor against cognitive decline.

We also observed gender differences. Positive associations between mental health and cognition were shown for females compared to males on the verbal memory tasks (immediate word recall delayed word recall) and males compared to females on the numerical tasks (numerical ability and subtraction). Previous studies reported gender differences in cognitive task performance, which appears to extend to the association between mental health and cognitive task performance. For example, a meta-analysis of a large sample of over 40,000 participants reported that females performed better than males in verbal recall tasks (Hirnstein, et al. 2023). Research supports a female advantage in memory related tasks, and some verbal subtests (e.g., California Verbal Learning Test, Boston Naming Test) show differences that are prevalent across the lifespan (23) although we note these differences are generally small. Past research has reported gender differences in mathematical performance too, but these have narrowed since the 1970s with males and females performing more similarly on numerical tasks in current times (24).

Our results showed a stronger association between mental health and the verbal memory tasks for married and divorced people compared to single people. Previous studies that used verbal recall tasks have also reported that singles were at greater risk of memory decline compared to married and divorced people (25). It appears that being married or living with others, either currently or in the past, has intrinsic qualitative cognitive elements that contribute to improved cognitive engagement and functioning. Regarding mental health, past research consistently reported that married people have the lowest prevalence of depression (26), also supported by the current results.

When we added social interaction to the regression model most of the results in the earlier models (i.e., self-reported cognitive ability, word recall tasks and numerical ability) persisted; however, there was no significant association found between social interaction and performance on subtraction and number series tasks. These results suggest that visiting friends had no or minimal effect on the association between mental health and cognition.

There have been longstanding difficulties with measuring the extent of social interaction in older adults. However, it is possible that the particular variable used in this study (a binary response scale) showed limited variability, and was therefore, not sufficiently sensitive to more nuanced individual differences in social isolation in older adults.

Studies have found an association between social isolation and poor cognition in people with depression or anxiety (27) suggesting that social networks and engagement in social activities may provide mental stimulation through interactions with others. Our results do not support that social interaction is more beneficial for cognitive abilities in older age. Instead, our results suggest that having good mental health is more beneficial as a protective factor against cognitive decline compared to social interaction. Perhaps having good mental health allows individuals to enjoy social interaction more and seek out opportunities for it, compared to those with poorer mental health. However, we often assume that having close social interaction and wider networks bring positive outcomes to the individual but social interaction may in some instances bring about negative outcomes. This might be particularly more prevalent to older adults for they might experience ageism and/or discrimination from family members and this might perpetuate negative stereotypes, thus negatively impacting mental health (Gordon, 2020).

### 4.1. Strengths and limitations

Our analyses are based on cognitive outcomes collected from a large population-based sample of over 7,000 respondents over 65 years of age providing high degree of statistical power and good scope for generalisability. Cognitive outcomes were collected at Wave 3 only, and therefore, our results are based on cross-sectional data, which offers a larger sample compared to a longitudinal approach. However, the study is based on correlational evidence only, and the cognitive tasks were limited. Repeated cognitive testing in subsequent waves will allow us to compare our sample across different waves, explore potential changes over time and rule out cohort effects; a disadvantage of cross-sectional data.

### 4.2. Conclusion

This study examined the relationship among mental health, cognitive abilities, and socio- demographic factors using a substantial representative population sample of over 7,000 older adults over 65 years old. We found that a positive relationship between mental health and cognitive abilities. This relationship was impacted by ethnicity, gender, education, and marital status.

## Data Availability

We obtained the data from the Understanding Society website (www.understandingsociety.ac.uk).

## Notes

### Competing Interest Statement

The authors have declared no competing interest.

### Clinical Trial

NA

### Funding Statement

The author(s) received no specific funding for this work.

## References

1. University of Essex. Understanding Society: Waves 1-13, 2009-2022 and Harmonised BHPS: Waves 1-18, 1991-2009: Special Licence Access. [data collection]. 17th Edition. UK Data Service. SN: 6931. 2023.

2. Aminzadeh F, Dalziel WB, Molnar FJ, Garcia LJ. Symbolic meaning of relocation to a residential care facility for persons with dementia. Aging & Mental Health. 2009;13(3):487–96. doi:10.1080/13607860802607314

3. Craik FI, Salthouse TA. The handbook of aging and cognition: Psychology press; 2011.

4. Chodosh J, Miller-Martinez D, Aneshensel CS, Wight RG, Karlamangla AS. Depressive symptoms, chronic diseases, and physical disabilities as predictors of cognitive functioning trajectories in older Americans. J Am Geriatr Soc. 2010;58(12):2350–7. doi: 10.1111/j.1532-5415.2010.03171.x

5. Lim MH, Eres R, Vasan S. Understanding loneliness in the twenty-first century: an update on correlates, risk factors, and potential solutions. Soc Psychiatry Psychiatr Epidemiol. 2020;55:793–810. doi:10.1007/s00127-020-01889-7

6. Zhou Z, Mao F, Zhang W, Towne SD, Jr., Wang P, Fang Y. the association between loneliness and cognitive impairment among older men and women in China: a nationwide longitudinal study. Int J Environ Res Public Health. 2019;16(16). doi:10.3390/ijerph16162877

7. Hertzog C, Kramer AF, Wilson RS, Lindenberger U. Enrichment effects on adult cognitive development: can the functional capacity of older adults be preserved and enhanced? Psychological Science in the Public Interest. 2008;9(1):1–65. 10.1111/j.1539-6053.2009.01034

8. Harada CN, Natelson Love MC, Triebel KL. Normal cognitive aging. Clin Geriatr Med. 2013;29(4):737-52. doi:10.1016/j.cger.2013.07.002

9. Gallagher D, Kiss A, Lanctot K, Herrmann N. Depressive symptoms and cognitive decline: a longitudinal analysis of potentially modifiable risk factors in community dwelling older adults. Journal of affective disorders. 2016;190:235–40. 10.1016/j.jad.2015.09.046

10. Birken M, Chipp B, Shah P, Olive RR, Nyikavaranda P, Hardy J, et al. Exploring the experiences of loneliness in adults with mental health problems: a participatory qualitative interview study. PLOS ONE, 18(3):e0280946. doi:10.1101/2022.03.02.22271346

11. Seshadri S, Wolf PA, Beiser A, Au R, McNulty K, White R, et al. Lifetime risk of dementia and Alzheimer’s disease. The impact of mortality on risk estimates in the Framingham Study. Neurology. 1997;49(6):1498–504. doi:10.1212/wnl.49.6.1498

12. Kuiper JS, Smidt N, Zuidema SU, Comijs HC, Oude Voshaar RC, Zuidersma M. A longitudinal study of the impact of social network size and loneliness on cognitive performance in depressed older adults. Aging & Mental Health. 2020;24(6):889–97. doi:10.1080/13607863.2019.1571012

13. Cohen-Mansfield J, Hazan H, Lerman Y, Shalom V. Correlates and predictors of loneliness in older-adults: a review of quantitative results informed by qualitative insights. Int Psychogeriatr. 2016;28(4):557–76. doi: 10.1017/S1041610215001532

14. DiNapoli EA, Wu B, Scogin F. Social isolation and cognitive function in Appalachian older adults. Research on Aging. 2014;36(2):161–79. doi:10.1177/0164027512470704

15. McFall SL, Garrington C. Understanding society: early findings from the first wave of the UK’s Household Longitudinal Study. University of Essex: Institute for Social and Economic Research; 2011.

16. Goldberg D, Williams P. General health questionnaire. Granada Learning Group London; 1988.

17. Lesage F-X, Martens-Resende S, Deschamps F, Berjot S. Validation of the General Health Questionnaire (GHQ-12) adapted to a work-related context. Open J Prev Med. 2011;1(02):44. doi: 10.4236/ojpm.2011.12007

18. Bauermeister S, Bunce D. Poorer mental health is associated with cognitive deficits in old age. Aging, Neuropsychology, and Cognition. 2015;22(1):95–105. doi:10.1080/13825585.2014.893554

19. Guo M, Wang Y, Carter K. Racial/ethnic and nativity differences in adversity profiles among middle-aged and older adults. Aging & mental health. 2024;28(2):319–29. 10.1080/13607863.2023.2251421

20. Díaz-Venegas C, Downer B, Langa KM, Wong R. Racial and ethnic differences in cognitive function among older adults in the USA. Int J Geriatr Psychiatry. 2016;31(9):1004–12. doi: 10.1002/gps.4410

21. Tari B, Künzi M, Pflanz CP, Raymont V, Bauermeister S. Education is power: preserving cognition in the UK biobank. Frontiers in Public Health. 2023;11:1244306. doi: 10.3389/fpubh.2023.1244306

22. Sperandei S, Page A, Spittal MJ, Pirkis J. Low education and mental health among older adults: the mediating role of employment and income. Soc Psychiatry Psychiatr Epidemiol. 2023;58(5):823–31. doi:10.1007/s00127-021-02149-y

23. Kheloui S, Jacmin-Park S, Larocque O, Kerr P, Rossi M, Cartier L, et al. Sex/gender differences in cognitive abilities. Neurosci Biobehav Rev. 2023:105333. doi: 10.1016/j.neubiorev.2023.105333

24. Hyde JS. Sex and cognition: gender and cognitive functions. Curr Opin Neurobiol. 2016;38:53-6. doi: 10.1016/j.conb.2016.02.007

25. Mousavi-Nasab S-M-H, Kormi-Nouri R, Sundström A, Nilsson LG. The effects of marital status on episodic and semantic memory in healthy middle-aged and old individuals. Scand J Psychol. 2012;53(1):1–8. doi: 10.1111/j.1467-9450.2011.00926.x

26. Bulloch AG, Williams JV, Lavorato DH, Patten SB. The depression and marital status relationship is modified by both age and gender. Journal of Affective Disorders. 2017;223:65–8. doi: 10.1016/j.jad.2017.06.007

27. Evans IE, Llewellyn DJ, Matthews FE, Woods RT, Brayne C, Clare L. Social isolation, cognitive reserve, and cognition in older people with depression and anxiety. Aging & Mental Health. 2019;23(12):1691–700. 10.1080/13607863.2018.1506742

